# Rapid and accurate detection of novel coronavirus SARS-CoV-2 using CRISPR-Cas3

**DOI:** 10.1101/2020.06.02.20119875

**Authors:** Kazuto Yoshimi, Kohei Takeshita, Seiya Yamayoshi, Satomi Shibumura, Yuko Yamauchi, Masaki Yamamoto, Hiroshi Yotsuyanagi, Yoshihiro Kawaoka, Tomoji Mashimo

## Abstract

Novel coronavirus SARS-CoV-2 outbreaks have rapidly spread to multiple countries, highlighting the urgent necessity for fast, sensitive, and specific diagnostic tools for virus surveillance. Here, the previously unknown collateral single-stranded DNA cleavage we observed with type I CRISPR-Cas3 highlights its potential for development as a Cas3-mediated rapid (within 40 min), low-cost, instrument-free detection method for SARS-CoV-2. This Cas3-based assay is comparable with Cas12- and real-time reverse-transcriptase PCR-based assays in its speed and sensitivity, but offers greater specificity for single-base-pair discrimination while negating the need for highly trained operators. These findings support the use of CRISPR diagnostics for point-of-care testing in patients with suspected SARS-CoV-2 infections.

---

The coronavirus disease 2019 (COVID-19) pandemic caused by the severe acute respiratory syndrome coronavirus 2 (SARS-CoV-2), which emerged in Wuhan, China in late December 2019, has resulted in more than 5.35 million laboratory-confirmed cases in over 210 countries, and more than 340,000 deaths as of May 25, 2020^1, 2^. The virus is mainly spread between people during close contact even before the symptoms appear^3^. During the COVID-19 outbreak on the Diamond Princess in Japan in February 2020, 3,700 passengers and crew members were kept aboard the ship for one month, mainly because of the capacity shortage of COVID-19 diagnostic kits^4^. Many countries use assays based on real-time (r) reverse-transcriptase (RT) polymerase chain reaction (PCR) (rRT-PCR) to detect viruses. However, the results of such assays on clinical samples from people with suspected SARS-CoV-2 infections are generally not ready until the day after sample collection because the samples need be shipped to reference laboratories for accurate diagnostic testing^5, 6^. rRT-PCR assays also require expensive equipment and well-trained personnel for their operation, making them unsuitable for point-of-care test (POCT) applications, but a POCT would have benefited the situation on the ship.

Enzymes from CRISPR-Cas systems have recently been adapted to rapidly, robustly, and sensitively detect viruses. These systems rely on Cas12a, otherwise called DETECTR (**D**NA **E**ndonuclease-**T**arg**E**ted **C**RISPR **T**rans **R**eporter)^7^, and Cas13, otherwise called SHERLOCK (**S**pecific **H**igh-sensitive **E**nzymatic **R**eporter Un**LOCK**ing)^8^. Both of these Cas enzymes, but not Cas9, exhibit nonspecific endonuclease activity in trans after binding to a specific cis target via programmable CRISPR RNAs (crRNAs). By combining isothermal amplification methods (e.g., **R**ecombinase **P**olymerase **A**mplification, RPA^9^; or Loop-mediated isothermal **AMP**lification, LAMP^10^) with reporting formats such as lateral flow detection with antigen-labeled reporters^11, 12^, DETECTR^12–14^ and SHERLOCK^15, 16^ have recently been used for rapid and highly sensitive SARS-CoV-2 detection. The Cas13-based strategy, PAC-MAN (**P**rophylactic **A**ntiviral **C**RISPR in hu**MAN** cells), has also been shown to inhibit and degrade SARS-CoV-2 viral RNA in respiratory epithelial cells^17^.

We and other groups have recently reported that Class 1 Type I-E CRISPR-Cas from *Escherichia coli*^18^ and *Thermobifida fusca*^19^, both of which employ a crRNA-bound multiprotein complex (the Cas complex) for antiviral defense (Cascade) and a Cas3 endonuclease, can mediate targeted DNA cleavage with long-range deletions in human cells. Despite type I-E *E. Coli* CRISPR being one of the most thoroughly biochemically characterized *in vitro* plasmid DNA degradation-inducing systems^20, 21^, whether the CRISPR-Cas3 system can mediate the target-activated, nonspecific single-stranded DNA (ssDNA) cleavage reported for Cas12 and Cas13^7, 8^remains an open question. Here, we report on the development of **C**as3-**O**perated **N**ucleic **A**cid detectio**N** (CONAN), an *in vitro* nucleic acid-detection platform (Figure 1a). When combined with isothermal amplification methods, CONAN provides a rapid, sensitive and instrument-free detection system for SARS-CoV-2 POCTs.

**Figure 1.**
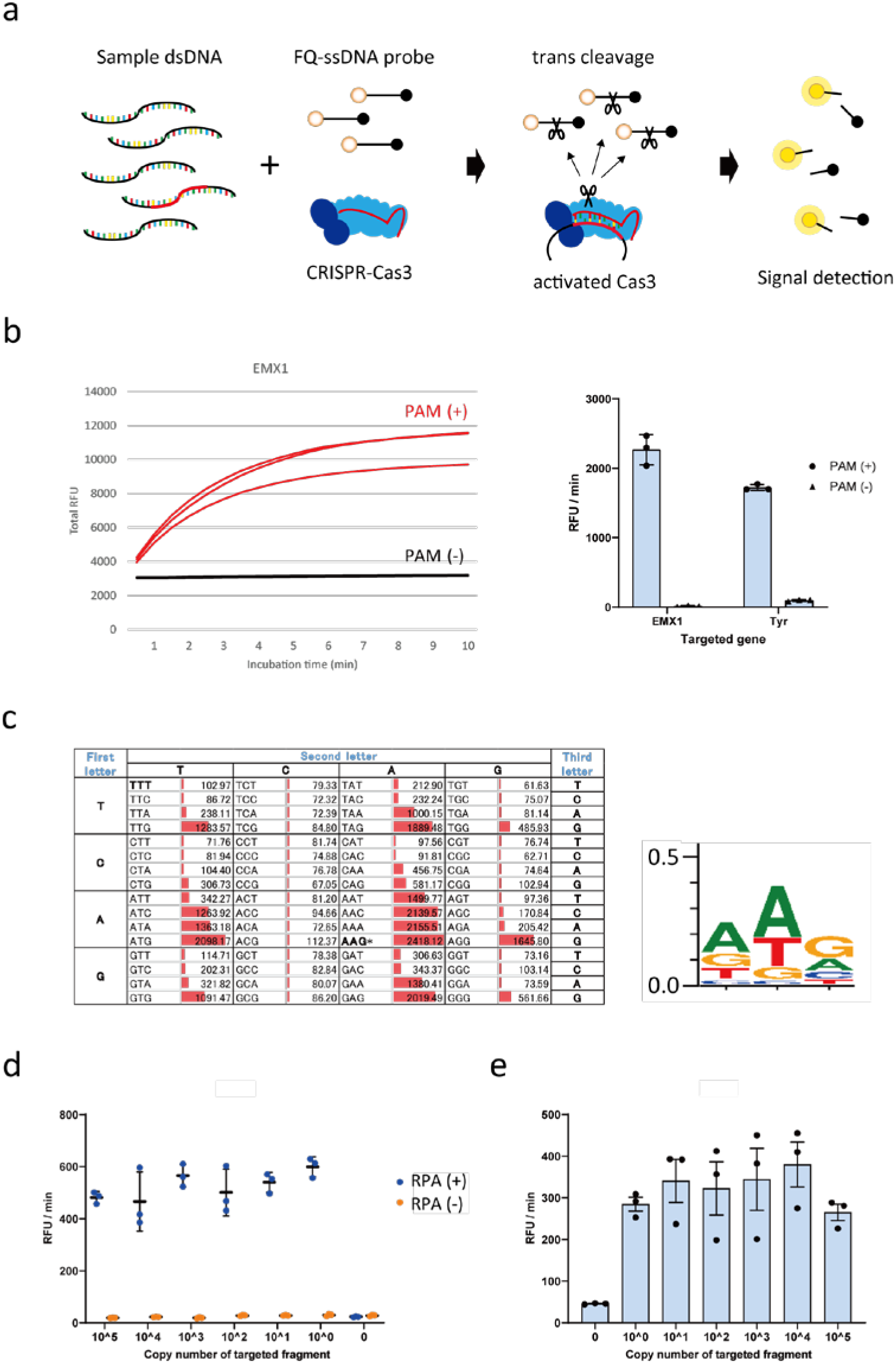
Cas3-operated nucleic acid detection (CONAN). **a**, Schematic representation of the CONAN *in vitro* nucleic acid-detection platform. The *E. coli* CRISPR-Cas3 complex contains Cas3, Cas5, Cas6, Cas7, Cas8, and Cas11 proteins and CRISPR RNA (crRNA). FQ-ssDNA, fluorophore and quencher-labeled single-stranded DNA probe. **b**, CRISPR-Cas3 mediates collateral ssDNA cleavage after targeting hEMX1-dsDNA (or mTyr-dsDNA) in fragments with PAMs (+; red), but not in fragments without PAMs (-, black). **c**, Screening for collateral activity on the 64 possible PAM target sites containing each of the three-nucleotide sequences. **d**, CONAN assay on isothermal RPA amplicon products detects a single copy of the activator fragments. **e**, CONAN RPA also detects a single-copy activator when mixed with the abundant mouse genomic DNA. RFU, relative fluorescence unit.

First, to investigate whether Class 1 CRISPR-Cas3 exhibits trans-cleavage activity on collateral ssDNA, we assembled the *E. Coli* Cas3 protein (20 ng/µL), Cascade with its crRNA (32 ng/µL), the double-stranded DNA (dsDNA) activator (2 ng/µL) for two target genes, human EMX1 or mouse Tyr (Table S1 and S2), and a fluorophore quencher (FQ)-labeled ssDNA probe (4.5 ng/µL) in the reaction buffer (5 mM HEPES-K pH7.5, 60 mM KCl, 10 mM MgCl_2_, 10 μM CoCl_2_, and 2.5 mM ATP). After 10 min incubation at 37ºC, we observed non-specific trans-ssDNA cleavage after site-specific dsDNA cutting within the positive (+) strand, but not the negative (-) strand (Figure 1b). We screened all 64 possible target sites containing each of the three-nucleotide protospacer adjacent motif (PAM) sequences (Figure 1c and Fig. S1a), and observed significant cleavage activities with 14 PAM types. The highest activity was from AAG, whereas no activity was seen with the first and the second C nucleotides and the third T nucleotide in the PAM. Therefore, we called this protocol Cas3-Operated Nucleic Acid Detection, or CONAN (Figure 1a).

We then investigated the detection sensitivity of CONAN by diluting the hEMX1 or mTyr dsDNA activator with Cas3, Cascade and FQ-ssDNA in the reaction buffer. CONAN’s limit of detection (LoD) was > 1.0 × 10^10^ copies for the activator (Fig. S1b). To improve the LoD, we performed isothermal RPA at 37ºC for 30 min, followed by a 10 min incubation with Cascade and Cas3, thereby determining the activator’s single-copy sensitivity level (Figure 1d). We also achieved robust detection of the single-copy activator after mixing it with abundant mouse genomic DNA (Figure 1e).

Detecting FQ-ssDNA cleavage needs a microplate reader for fluorescence intensity measurement. Instead of this laboratory instrument, a lateral flow strip can be used for instrument-free and portable diagnosis by the virus POCT^11, 12^. In principle, abundant reporter accumulates anti-FITC antibody-gold nanoparticle conjugates at the first line (negative) on the strip, while cleavage of the reporter would reduce accumulation on the first line and result in signal on the second line (positive) with < 2 min of flow (Fig. S2). Using this lateral flow strip we achieved a one-pot assay with CONAN-RPA, thereby detecting a single copy level of the target dsDNA within 40 min.

We next examined whether the CONAN lateral flow assay would be effective for SARS-CoV-2 diagnosis (Figure 2a), as has been recently reported for DETECTR^12–14^ and for SHERLOCK^15, 16^. We designed primers to amplify the N (nucleoprotein) gene regions (N1 and N2) from SARS-CoV-2 (Table S3), which overlap with the region used in the DTECTER-based assay^13^, along with primers for the qRT-PCR assay from the US CDC^22^. The qRT-PCR assay successfully amplified both the N1 and N2 regions of SARS-CoV-2, with LoDs of < 10^2^ copies (Figure 2b). We compared the CONAN- and DETECTR-based assays by designing crRNAs in the N1 and N2 regions (Table S1). However, RT-RPA followed by Cas3-based CONAN or Cas12-based DETECTOR in the one-step 37ºC 1-h incubation did not effectively detect SARS-CoV-2, probably because the N1 and N2 primers we designed did not match the sensitivity of the RT-based assay (Fig. S3a).

**Figure 2.**
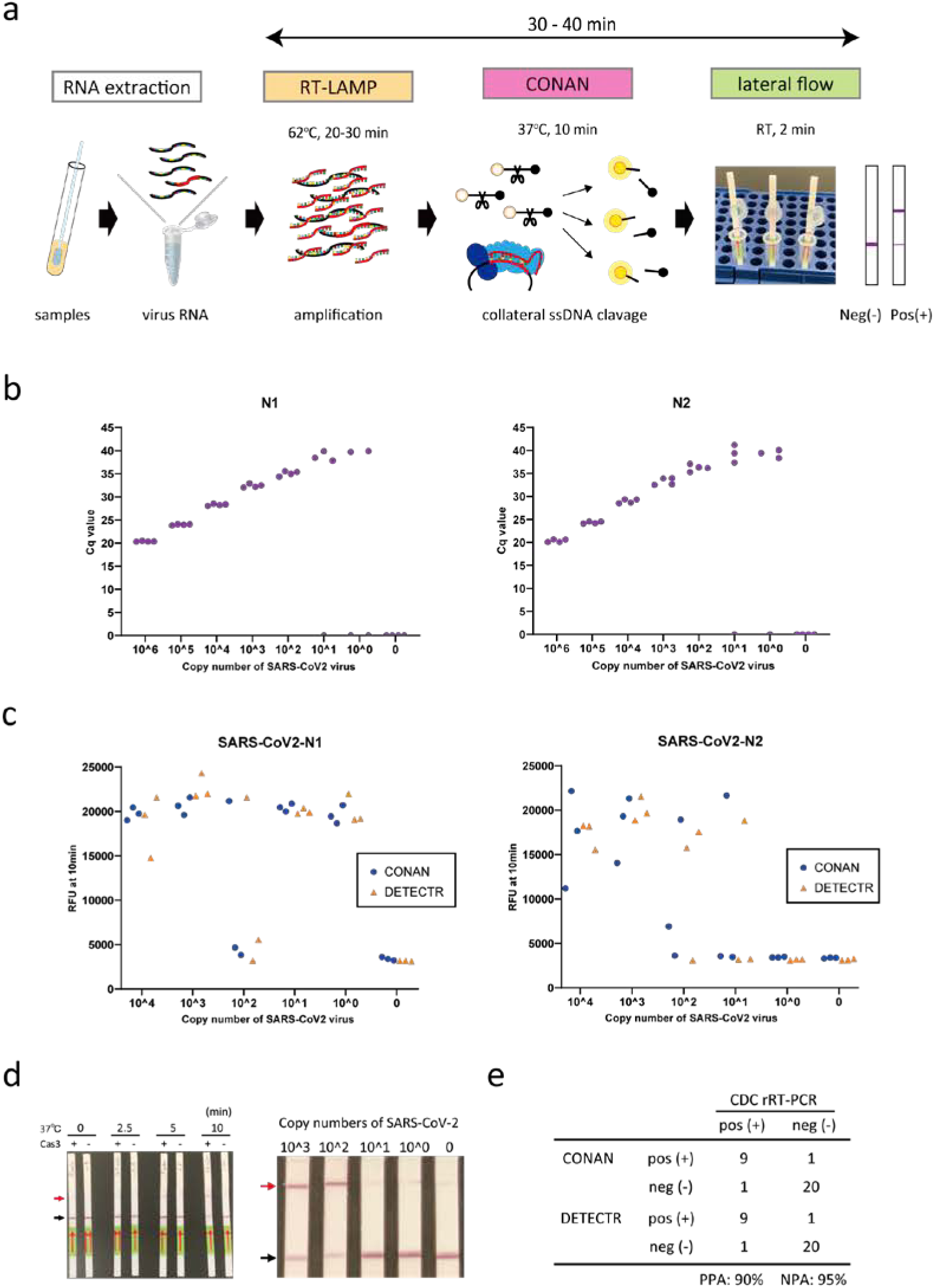
CRISPR-Cas3-based assay for rapid and sensitive detection of SARS-CoV-2. **a**, Schematic representation of the CONAN SARS-CoV-2 detection assay including a conventional RNA extraction step, RT-LAMP (62ºC, 20–30 min), CONAN (37ºC, 10 min), and lateral flow (RT, 2 min). **b**, The limit of detection (LoD) of the US CDC’s rRT-PCR assay amplification of the N1 and N2 regions of the SARS-CoV-2 N gene. **c**, LoD of CONAN-based and DETECTR-based assays for the N1 and N2 region of SARS-CoV-2. **d**, LoD of the CONAN-based lateral flow assay for the N1 region of SARS-CoV-2. Cq: cycle quantification value. RFU, relative fluorescence unit. **e**, Comparison of the SARS-CoV-2 CONAN and DETECTR assays using 31 clinical samples (10 positive and 21 negative for COVID-19 by the CDC rRT-PCR assay). PPA: positive predictive agreement. NPA: negative predictive agreement.

Designing primers for LAMP assays can be complicated, but the upside is that these assays seem to be less sensitive to inhibitors or off-target nucleic acid contamination in the samples. RT-LAMP at 62ºC for 30 min, followed by CONAN or DETECTR for 10 min at 37ºC, both specifically detected SARS-CoV-2 with crRNA–N1 and –N2 (Figure 2c). The LoDs for CONAN-LAMP and DETECTR-LAMP (< 10^2^ copies) compare favorably with the CDC qRT-PCR assay for SARS-CoV-2 detection^5, 13, 23^. This rapid detection by CONAN-LAMP was achieved at 62ºC for 30 min, with the lateral flow strip achieving a LoD of < 10^2^ copies for SARS-CoV-2 (Figure 2d and Supplementary Video).

Finally, we tested the extracted RNA samples from 10 PCR-positive COVID-19 patients and 15 PCR-negative samples from nasopharyngeal swabs, using the CONAN RT-LAMP and DETCTR RT-LAMP assays with lateral flow strip readouts (Figure 2e). SARS-CoV-2 was detected by the CONAN RT-LAMP assay in 9 of 10 patient swabs and detected in one of 21 negative swab samples (positive predictive agreement; 90% and negative predictive agreement; 95%). One negative swabs from COVID-19 patients was confirmed to be below the established LoD of < 10^2^ copies (Fig. S3b). This 94% detection rate (29/31) is comparable with that of the DETECTR RT-LAMP assay in this study (Figure 2e) and that previously reported for a COVID-19 POCT^13^.

CRISPR-based diagnostic (CRISPR-dx) nucleic acid tests have been used to detect the presence DNA or RNA in a range of samples across biotechnology and health areas^7, 8, 24, 25^. They mainly include type V Cas12a DETECTR^12–14^ and Cas13 type VI SHERLOCK^15, 16^, which both belong to Class II CRISPR. Here, we report on the discovery of Class I Cas3-mediated collateral trans cleavage activity. Our newly developed a CONAN lateral flow assay, which uses this type of collateral cleavage, facilitated the rapid, robust, and sensitive detection of novel coronavirus SARS-CoV-2. Unlike the single Cas12a and Cas13 platforms, CONAN needs multiple Cas proteins (Cas3, 5, 6, 7, 8, and 11), but they are stable when premixed and stored at around 4ºC. Interestingly, unlike DETECTR, CONAN shows high specificity for single-base-pair discrimination within the PAM site (Fig. S4), supporting the applicability of CONAN-based detection assays for POCTs, even with novel emerging coronavirus mutants.

Several other SARS-CoV-2 detection assays and their protocols have been published on the World Health Organization’s website or are currently under development^26^. For SARS-CoV-2-based tests, rapid collection of appropriate specimens from patients is a priority goal for clinical management and outbreak control. The RT-PCR-based assays, despite their established efficiency and accuracy for SARS-CoV-2 detection, require highly specialized personnel and expensive equipment for implementation. In contrast, serology IgM/IgG tests with lateral flow immunoassays can rapidly and sensitively determine the infection rate in a population, although IgG antibodies to SARS-CoV-2 are generally only detectable 10–14 days post-infection. SARS-CoV-2 antigen tests can directly detect viral components without the amplification steps needed for RT-PCR and LAMP; however, the technology is new and the evidence for its accuracy in coronavirus diagnosis is still being evaluated. CRISPR-dx is also cheaper and allows portable diagnostic tests for the assessment of suspected cases without the need for sophisticated equipment. Although still awaiting evaluation, CRISPR-dx methods have high potential for transforming pathogen identification and other aspects of infectious disease diagnostics, such as that relating to SARS-CoV-2.

## Methods

### CRISPR preparation

CRISPR-Cas3 and CRISPR-Cas12a target sites were based on the human EMX1 gene, the mouse Tyr gene, and the N gene from SARS-CoV-2 (Supplementary Table 1). Cas3 and Cascade proteins from *E. coli* and the crRNA complex (EcoCascade-crRNA) were produced with reference to a previous report, with several modifications^27^. Briefly, the expressed EcoCas3 protein was purified using nickel affinity resin (Ni-NTA, QIAGEN, Venlo, Netherlands) and size-exclusion chromatography (Superdex 200 Increase 10/300 GL; Thermo Fisher Scientific, Waltham, Massachusetts) in 200 mM NaCl, 10% Expressed recombinant Cascade-crRNA was also purified using Ni-NTA resin and size-exclusion glycerol, 1mM DTT and 20 mM HEPES-Na (pH 7.0). Expressed recombinant Cascade-crRNA was also purified using Ni-NTA resin and size-exclusion chromatography in 350 mM NaCl, 1mM DTT and 20 mM HEPES-Na (pH 7.0). AsCas12a and LbCas12a were purchased from Integrated DNA Technologies (IDT, Coralville, IA) and New England Biolabs (NEB, Ipswich, Massachusetts), respectively. Target-specific crRNAs were also purchased from IDT.

### DNA and RNA preparation

For the CRISPR-Cas3 activator templates, 60 bp fragments of hEMX1 and mTyr (which included a target site) were designed and purchased from IDT. PAM variants in hEMX1 were also designed to examine the correlation between collateral cleavage activity and PAM specificity (Supplementary Table 2). Total mouse genomic DNA from a C57BL/6 strain was used after purification (Maxwell RSC Cell DNA Purification Kit; Promega, Madison, Wisconsin). LAMP SARS-CoV-2 primers were designed against regions of the N gene using PrimerExplorer v.5 (Eiken Chemical Co.; https://primerexplorer.jp/). The primers used for isothermal PCR are listed in Supplementary Table 3.

Viral RNAs from SARS-CoV-2 were prepared according to the established protocol from the National Institute of Infectious Diseases in Japan^28^. Viral RNAs were purified from an infected TMPRSS2-expressing VeroE6 cell line using the QIAamp Viral RNA Mini Kit (QIAGEN) according to the manufacturer’s protocol. VeroE6/TMPRSS2 cells are available from the Japanese Collection of Research Bioresources Cell Bank in Japan (https://cellbank.nibiohn.go.jp/english/) (JCRB no. JCRB1819).

### Real-time rRT-PCR

Real-time rRT-PCR was used to determine SARS-CoV-2 RNA copy numbers using Reliance One-Step Multiplex RT-qPCR Supermix and CFX Connect (Bio-Rad Laboratories, Hercules, CA) according to the manufacturer’s protocols. N gene-specific primer and probe sets for real-time rRT-PCR assays and the SARS-CoV-2 plasmid (positive control) were purchased from IDT.

### CONAN assay

To characterize the Cas3 collateral cleavage assays, DNA templates were added to 100 nM Cascade-crRNA complex, 250 nM Cas3 and 2.5 mM ATP in CRISPR-Cas3 working buffer (60 mM KCl, 10 mM MgCl_2_, 50uM CoCl_2_, 5 mM HEPES-KOH pH 7.5), as previously described^27^. The ssDNA reporter probe (5’-/5HEX/AAGGTCGGA/ZEN/GTCAACGGATTTGGTC/3IBFQ/-3’) (250 nM) was added, and the probe’s cleavage-related change in the fluorescence signal was measured every 30 s for 10 min under 37°C incubation.

To detect DNAs, isothermal amplification and RPA were performed using the TwistAmp Basic kit (TwistDx, Maidenhead, UK) according to the manufacturer’s protocol. Template DNAs were amplified by incubation at 37°C for 20 min. To detect RNAs, isothermal amplification by the RT-LAMP method was performed using the WarmStart LAMP Kit (NEB) according to the manufacturer’s protocol. Template RNAs were reverse transcribed and amplified by incubation at 62 °C for 20 min. To detect low-copy-number molecules in the patients’ samples, the incubation time was extended to 30 min.

The CRISPR-Cas3 reaction mixture used for the CONAN method contained 100 nM EcoCascade-crRNA complex, 400 nM Cas3 and 2.5 mM ATP in the working buffer. AsCas12a and LbCas12a were prepared as described previously^13^ for use as positive controls for trans cleavage activity. Cas12a (50 nM) was incubated with 62.5 nM of crRNA in 1× NEBuffer 2.1 for 30 min at 37 °C. After amplification, 2 µl of the amplicon was combined with 18 µl of Cas3 and the Cascade-crRNA complex or the Cas12a-crRNA complex, and 250 nM of the ssDNA reporter probe was added. The fluorescence signal was measured every 30 sec at 37°C.

### Lateral flow assay

To optimize the CONAN assay for lateral flow readouts, the CRISPR-Cas3 reaction mixture was added to 200 nM of the Cascade-crRNA complex, with 400 nM Cas3 and 2.5 mM ATP in the working buffer. A 2 µl aliquot of the amplicon was added to 18 µl of Cas3, Cascade-crRNA complex and 500 nM of the ssDNA reporter probe (5’-/5-FITC/TAGCATGTCA/3-Biotin/-3’). The mixture was incubated for 10 min at 37 °C. After adding 50 µl of nuclease-free water, a lateral flow strip (Milenia HybriDetect 1; TwistDx) was added to the reaction tube and the result was visualized after approximately 2 min. A lower band close to the sample pad indicated a negative result (uncut probes), whereas an upper band close to the top of the strip indicated the 5′ end of the cut probes. Emergence of an upper band indicated a positive result (Fig. S2).

### Collection of human clinical samples

Clinical nasopharyngeal and oropharyngeal swab samples from patients infected with SARS-CoV-2 were collected by IMSUT hospital, according to protocols that were approved by the Research Ethics Review Committee of the Institute of Medical Science, the University of Tokyo. Negative nasopharyngeal swabs were collected from healthy donors at IMSUT. RNA from the samples of patients and healthy donors was extracted as described in the NIID-approved protocol (input 140 µl; elution, 60 µl) using the Viral RNA Mini kit (QIAGEN).

### Data availability

The data that support the findings of this study are available from the corresponding authors upon reasonable request.

## Acknowledgments

We thank Y. Kunihiro, T. Omoto and S. Kobori at Osaka University and M. Hoshi and A. Fukui at Tokyo University and S. Saji, S. Yamamoto, M. Omatsu and N, Godai at the RIKEN SPring-8 Center for their technical assistance. This project was supported in part by JSPS KAKENHI Grant Number 18H03974 (T.M.) and 19K16025 (K.Y.). Partially support also came from the Platform Project for Supporting Drug Discovery and Life Science Research (Basis for Supporting Innovative Drug Discovery and Life Science Research (BINDS)) from AMED under Grant Number JP20am0101070 (support numbers 1251 and 2463), from Research Program on Emerging and Re-emerging Infectious Diseases from AMED (JP19fk0108113), and from the National Institutes of Allergy and Infectious Diseases funded Center for Research on Influenza Pathogenesis (CRIP; HHSN272201400008C). We thank Sandra Cheesman, PhD, from Edanz Group (https://www.edanzediting.com/ac) for editing a draft of this manuscript.

## Author Contributions

K.Y. designed and performed most of the experiments and analyzed the data with assistance from S.S. and Y.Y. K.T. and M.Y. prepared the CRISPR-Cas proteins and performed the experiments. S.Y. H.Y. and. Y.K. prepared the clinical samples for CRISPR diagnostic testing. T.M. conceived and supervised the study, prepared the figures and wrote the manuscript with editorial contributions from all the authors.

## Conflict of interest

S.S. is employee of C4U. K.Y. and T.M. are co-founders and scientific advisors for C4U. The other authors declare no competing interests.

## Supplementary Information

**Supplementary Table 1.**
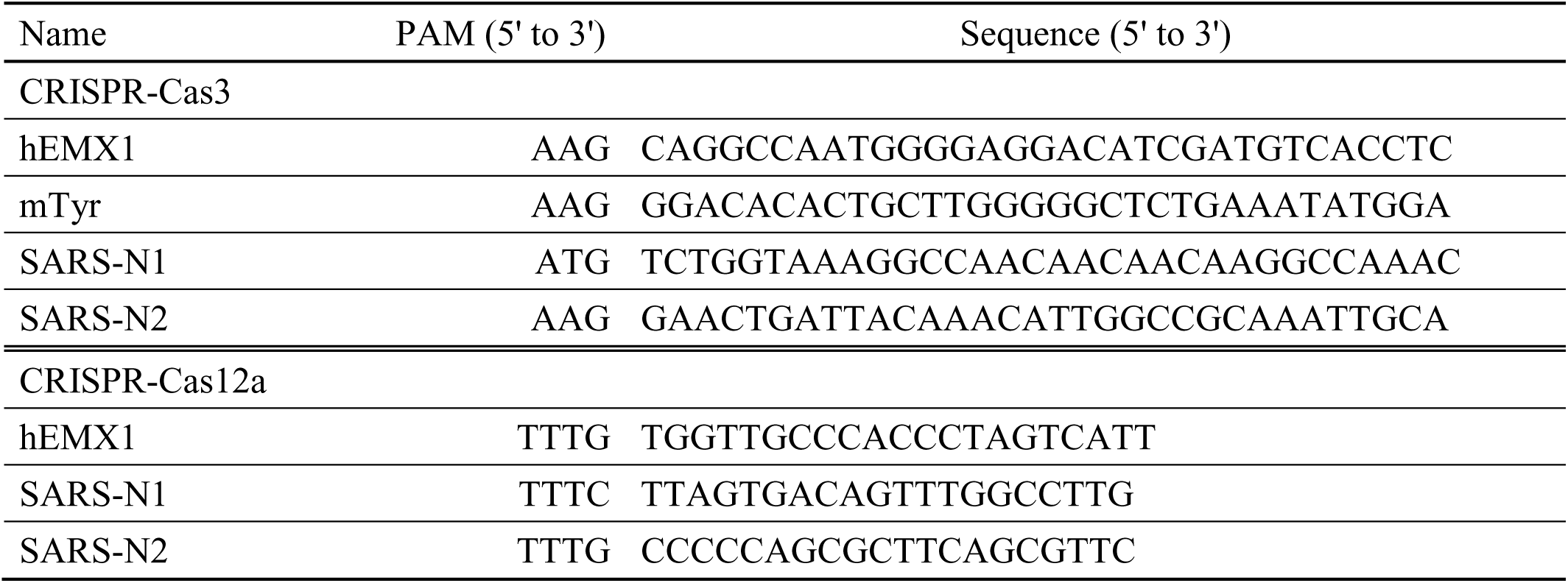
Target sequences for trans cleavage assay.

**Supplementary Table 2.**
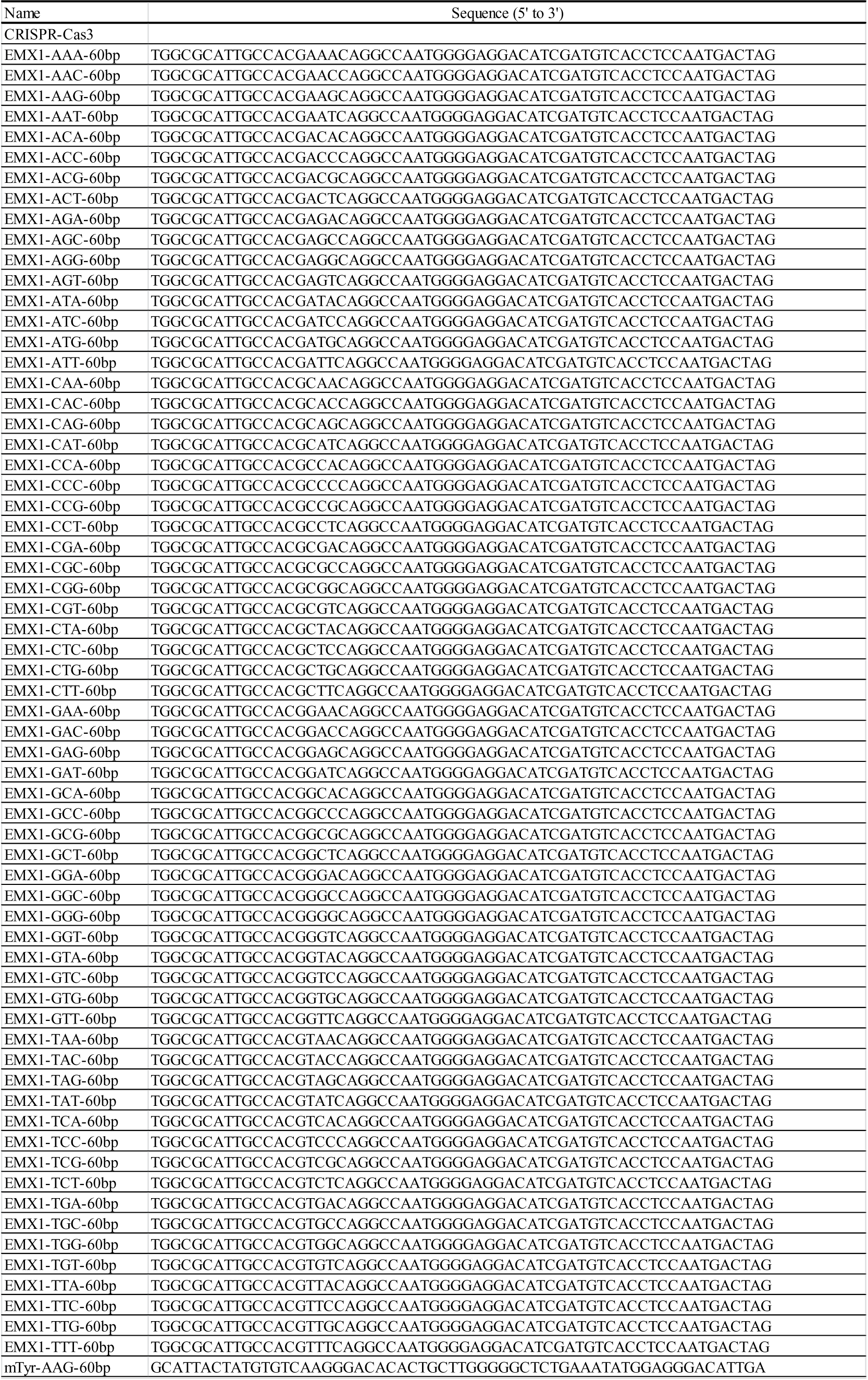
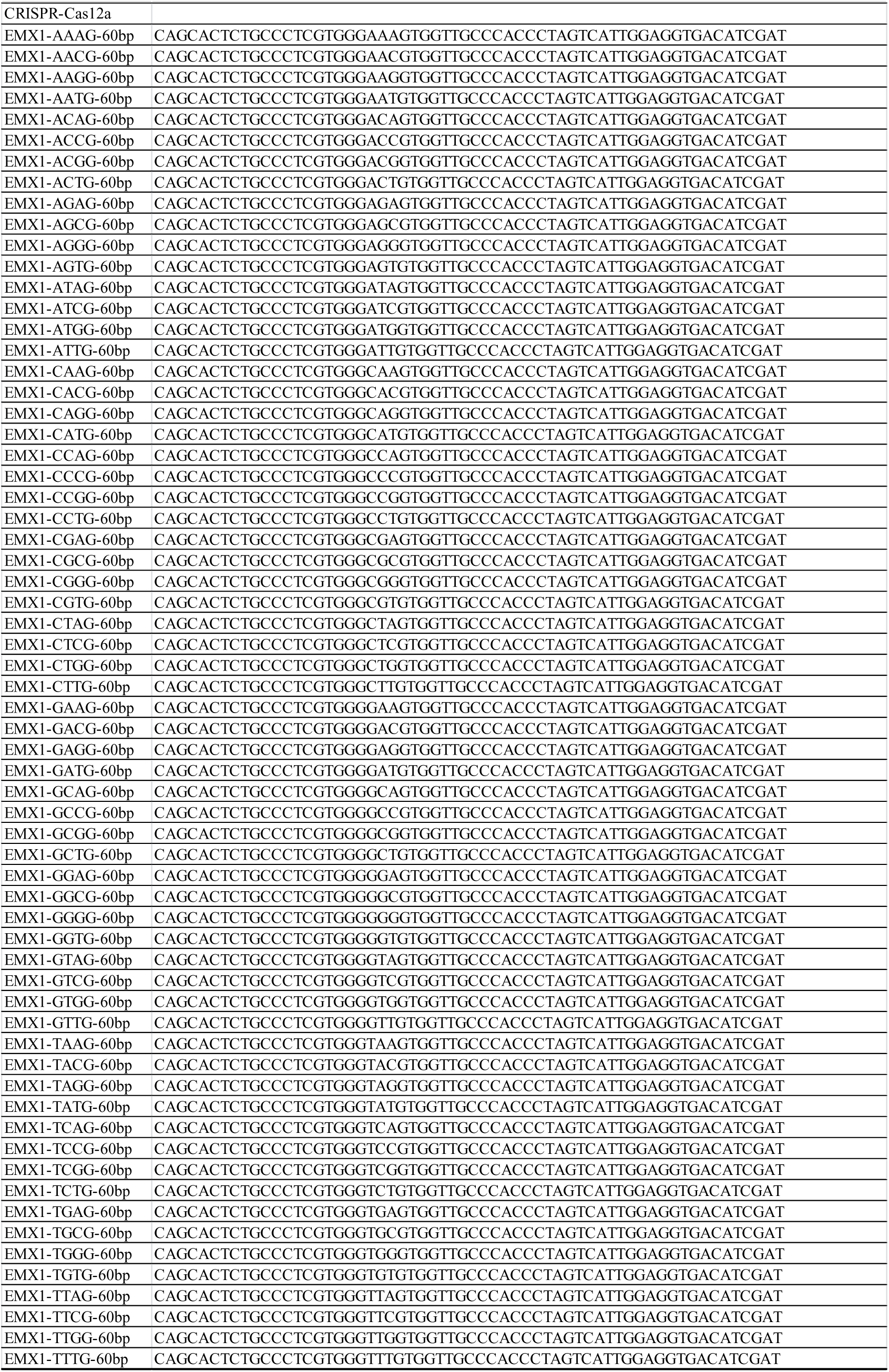
DNA fragment sequences for estimating PAM spesificity.

**Supplementary Table 3.**
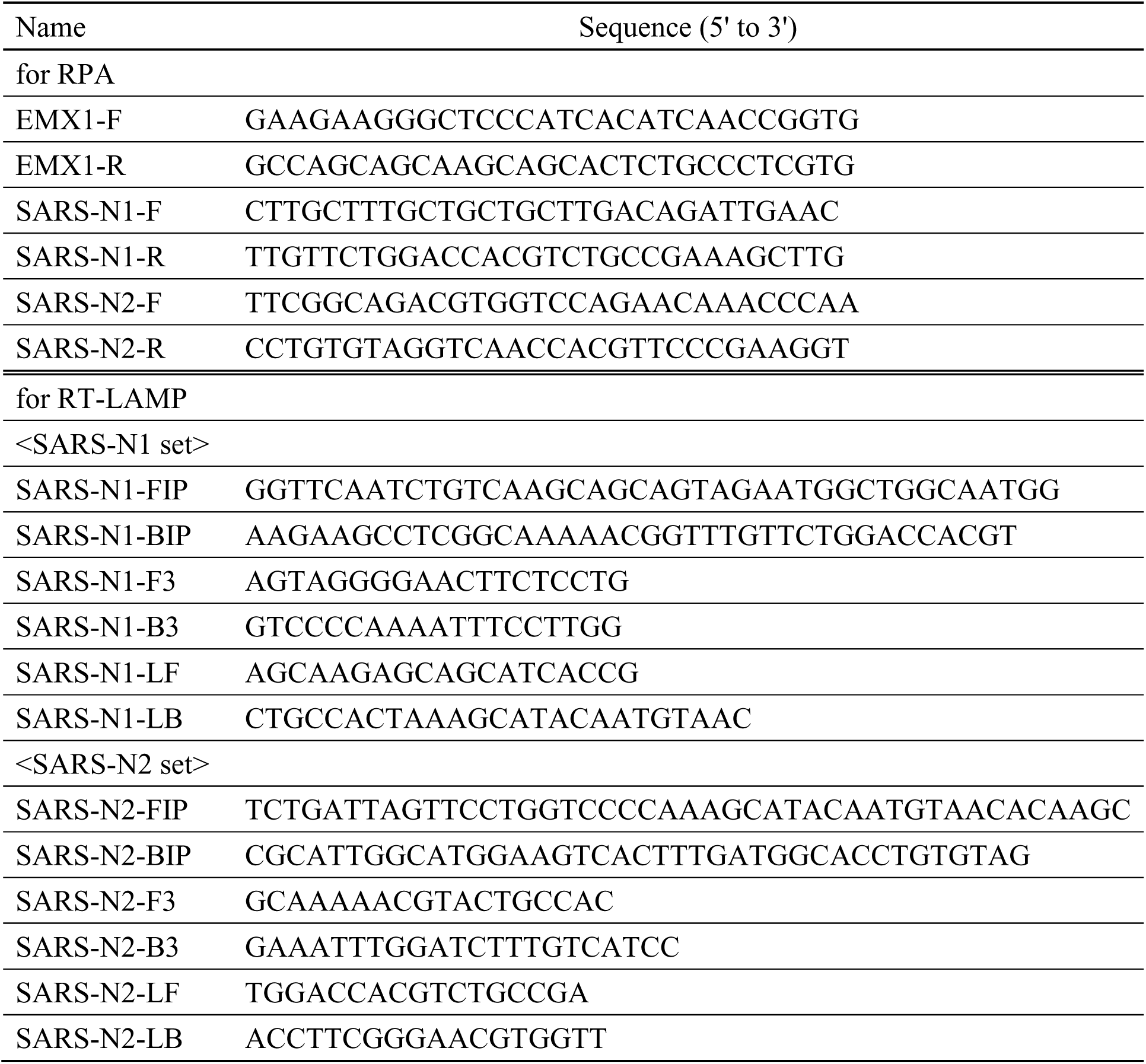
Primer sequences for isothermal PCR.

**Supplementary Figure 1.**
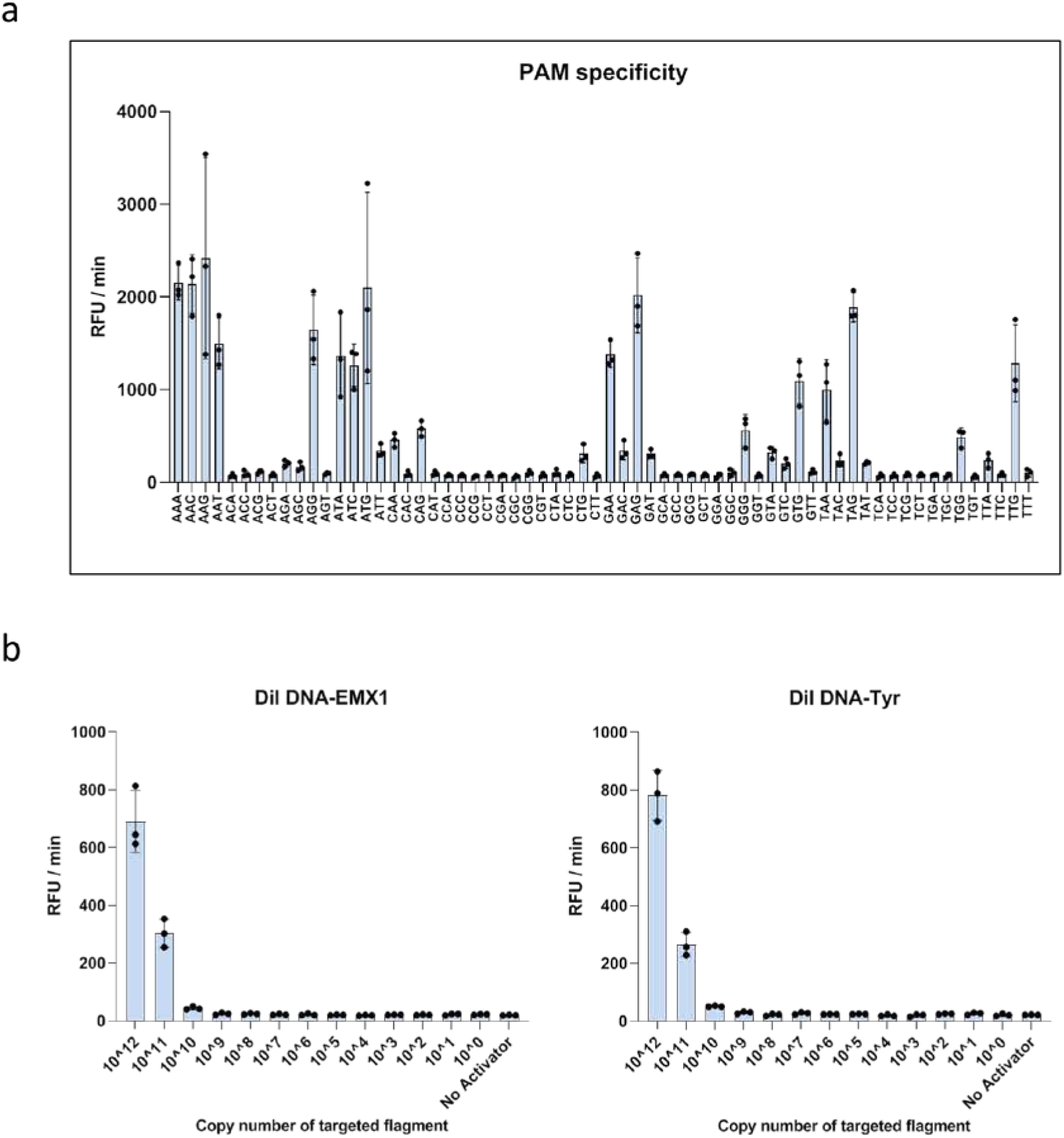
Cas3-operated nucleic acid detection (CONAN). **a**, CONAN assay screening of the 64 possible PAM target sites containing each of the three-nucleotide sequences (also see **Figure 1c**). **b**, Limit of detection (LoD) of the CONAN assay as estimated by dilution of the dsDNA activators hEMX1 or mTyr with Cas3, Cascade and FQ-ssDNA in the reaction buffer.

**Supplementary Figure 2.**
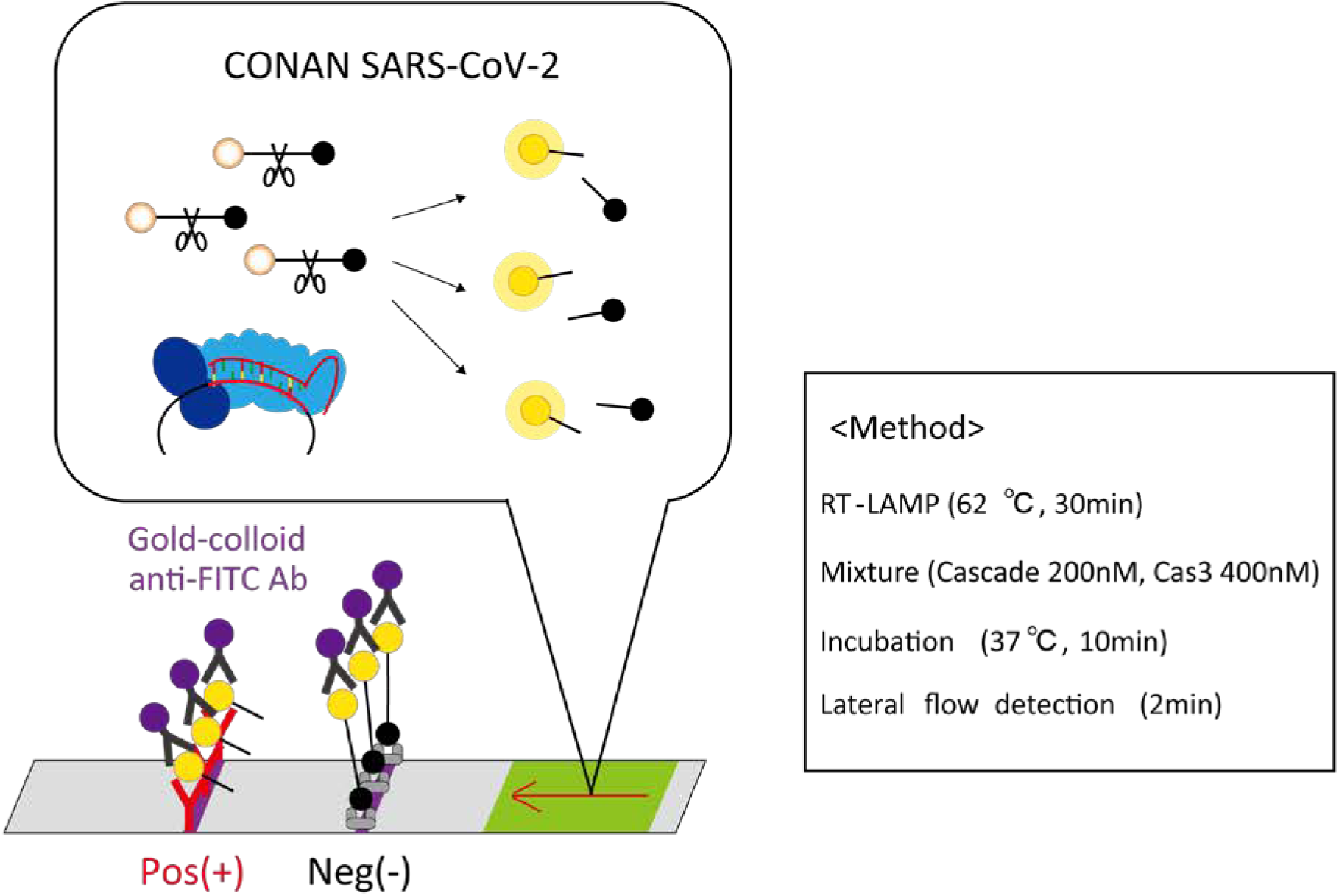
Lateral-flow detection with CONAN. Abundant uncleaved FQ-labeled ssDNA reporter accumulates anti-FITC antibody-gold nanoparticle conjugates on the first line (negative) of the strip, while the cleaved reporter reduces the accumulation at the first line, resulting in a signal on the second line (positive) with a < 2 min flow time.

**Supplementary Figure 3.**
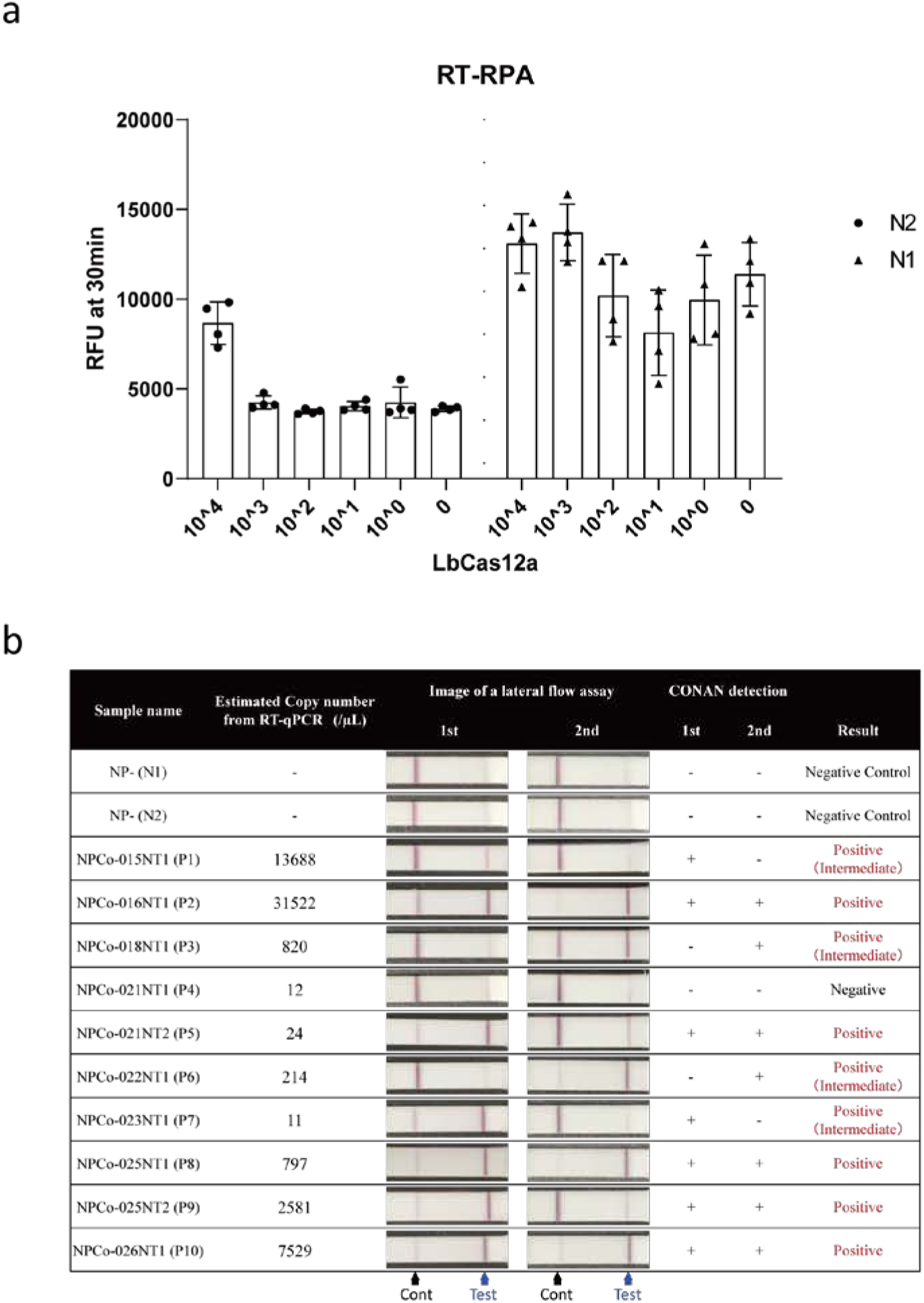
**a,** Limit of detection of the Cas12a-based DETECTR RT-RPA assay using N1 and N2 primers designed for the N gene region of SARS-CoV-2. **b**, SARS-CoV-2 CONAN lateral flow assays on 10 PCR-positive samples.

**Supplementary Figure 4.**
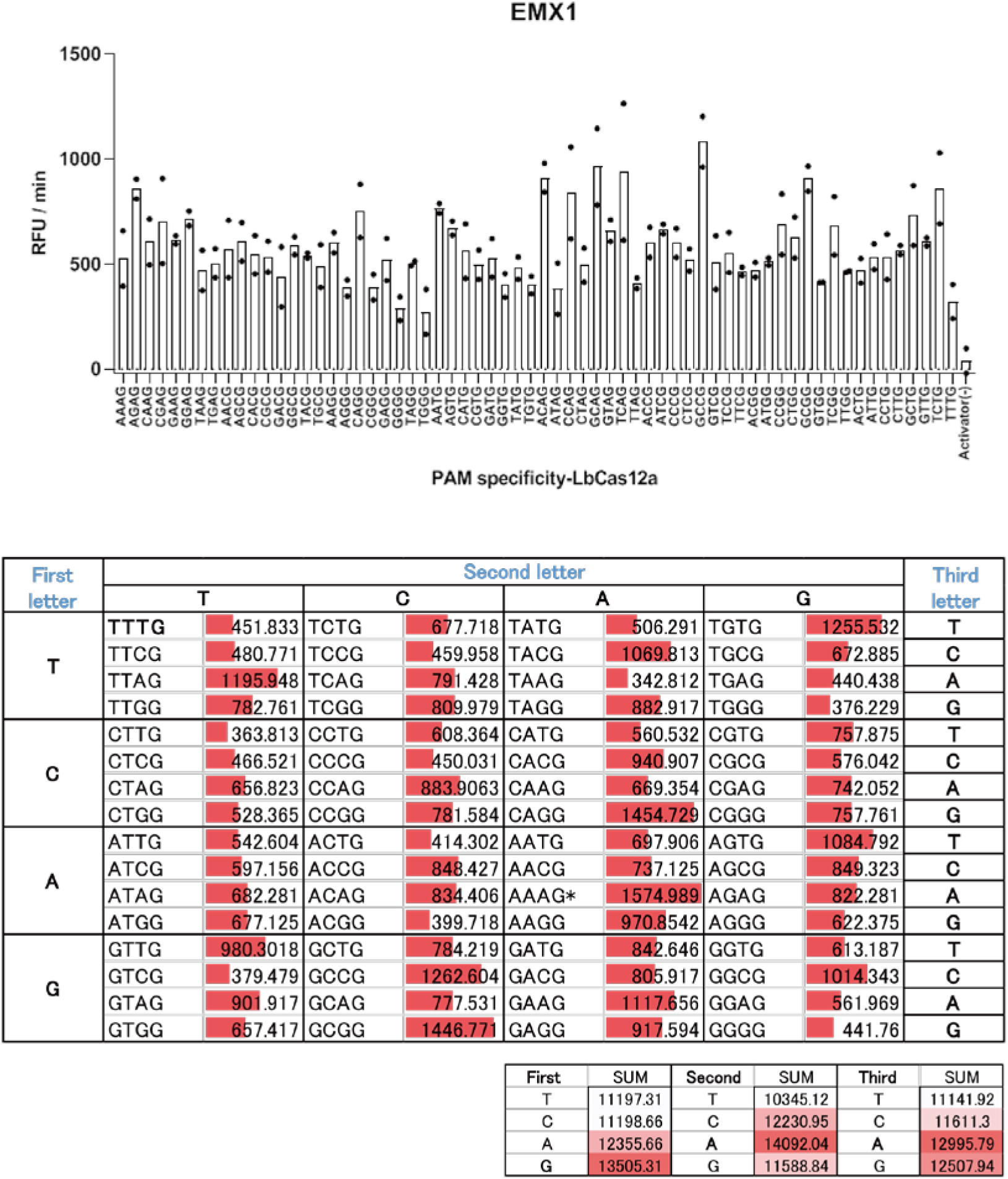
Cas12a-based DETECTR assay screening of the 64 possible target sites containing each of the three-nucleotide PAM sequences.

## Notes

### Author Declarations

Clinical nasopharyngeal and oropharyngeal swab samples from patients infected with SARS-CoV-2 were collected by IMSUT hospital, according to protocols that were approved by the Research Ethics Review Committee of the Institute of Medical Science, the University of Tokyo.

